# Assessing the effects of data drift on the performance of machine learning models used in clinical sepsis prediction

**DOI:** 10.1101/2022.06.06.22276062

**Authors:** Keyvan Rahmani, Rahul Thapa, Peiling Tsou, Satish Casie Chetty, Gina Barnes, Carson Lam, Chak Foon Tso

**Affiliations:** Dascena, Inc., 12333 Sowden Rd Ste B PMB 65148, Houston, Texas 77080-2059

**Keywords:** Data drift, sepsis, artificial intelligence, clinical decision support

## Abstract

**Background:** Data drift can negatively impact the performance of machine learning algorithms (MLAs) that were trained on historical data. As such, MLAs should be continuously monitored and tuned to overcome the systematic changes that occur in the distribution of data. In this paper, we study the extent of data drift and provide insights about its characteristics for sepsis onset prediction. This study will help elucidate the nature of data drift for prediction of sepsis and similar diseases. This may aid with the development of more effective patient monitoring systems that can stratify risk for dynamic disease states in hospitals.

**Methods:** We devise a series of simulations that measure the effects of data drift in patients with sepsis. We simulate multiple scenarios in which data drift may occur, namely the change in the distribution of the predictor variables (covariate shift), the change in the statistical relationship between the predictors and the target (concept shift), and the occurrence of a major healthcare event (major event) such as the COVID-19 pandemic. We measure the impact of data drift on model performances, identify the circumstances that necessitate model retraining, and compare the effects of different retraining methodologies and model architecture on the outcomes. We present the results for two different MLAs, eXtreme Gradient Boosting (XGB) and Recurrent Neural Network (RNN).

**Results:** Our results show that the properly retrained XGB models outperform the baseline models in all simulation scenarios, hence signifying the existence of data drift. In the major event scenario, the area under the receiver operating characteristic curve (AUROC) at the end of the simulation period is 0.811 for the baseline XGB model and 0.868 for the retrained XGB model. In the covariate shift scenario, the AUROC at the end of the simulation period for the baseline and retrained XGB models is 0.853 and 0.874 respectively. In the concept shift scenario and under the mixed labeling method, the retrained XGB models perform worse than the baseline model for most simulation steps. However, under the full relabeling method, the AUROC at the end of the simulation period for the baseline and retrained XGB models is 0.852 and 0.877 respectively. The results for the RNN models were mixed, suggesting that retraining based on a fixed network architecture may be inadequate for an RNN. We also present the results in the form of other performance metrics such as the ratio of observed to expected probabilities (calibration) and the normalized rate of positive predictive values (PPV) by prevalence, referred to as lift, at a sensitivity of 0.8.

**Conclusion:** Our simulations reveal that retraining periods of a couple of months or using several thousand patients are likely to be adequate to monitor machine learning models that predict sepsis. This indicates that a machine learning system for sepsis prediction will probably need less infrastructure for performance monitoring and retraining compared to other applications in which data drift is more frequent and continuous. Our results also show that in the event of a concept shift, a full overhaul of the sepsis prediction model may be necessary because it indicates a discrete change in the definition of sepsis labels, and mixing the labels for the sake of incremental training may not produce the desired results.

## 1. Introduction

In most application domains, the predictive power of machine learning (ML) degrades with time. This can be attributed to the fact that most ML models are built using historical datasets. This does not account for systematic changes that occur both to the distribution of the predictors and to the relationship between the predictors and the targets in future, which gradually degrade the performance of models trained on the historical data. This phenomenon is called “data drift”, or “data shift”.^1–3^ It can be mathematically represented by changes to the joint distribution of P(X, Y), where X represents the predictors and Y represents the targets of the machine learning algorithm (MLA).

Changes to P(X, Y) may occur as a result of changes in either of P(X), P(Y) or P(Y|X). These different changes do not have consistently used names. Rather, they are usually given different names by the authors. Jose G. Moreno-Torres provides a more unified naming convention for them by calling the changes to P(X) a covariate shift, to P(Y) a prior probability shift, and to P(Y|X) a concept shift.^1^ As per Moreno-Torres, a covariate shift is a change to the distribution of the predictors (i.e., input features); a prior probability shift is a change to the distribution of the target variables; a concept shift is the change to the statistical relationship P(Y|X) between predictors and targets. We will apply these terms within this paper with only one exception that uses the terms prevalence and prior probability shift interchangeably, as the use of the term prevalence is much more common in the medical community than prior probability.

In sepsis prediction, concept drift would refer to a change in the definition of sepsis (concept) over time. Such a change would be explicitly known, as it would normally follow any changes to the gold standard definition for sepsis diagnosis. Though these changes are not typically gradual, they are sometimes dependent upon the healthcare providers’ interpretation of these definitions, which may change over time.^4,5^ We hypothesize that concept changes occur explicitly with a marked transition time. On the other hand, the covariate shift in clinical applications can be implicit and can potentially happen gradually.^6^ In extenuating circumstances however, such as a global pandemic, changes in both covariates and prevalence may occur suddenly. The COVID-19 pandemic is an example of how such a sudden change can cause a drastic impact within hospitals and other healthcare settings.^7–11^

In this paper, we try to understand which categories of data drift (covariate shift, concept shift, prior probability/prevalence shift) are more likely to happen in regard to the condition of sepsis. We provide insights about the characteristics of data drift for sepsis prediction by devising a series of simulations that measure data drift in a sepsis population. The results of this study may enable healthcare providers to better understand the nature of the data drift for sepsis and similar diseases, which may ultimately aid with the development of more effective patient monitoring systems that can stratify risk for dynamic disease states. Sepsis is a serious condition that develops as a result of the body’s extreme response to an infection.^12^ It is considered a global health priority by the World Health Organization (WHO),^12^ and in the U.S. alone, it imposes a $20B cost on the healthcare system^13^ and affects 750,000 patients annually.^14^

Although the subject of data drift in ML in general is not new^1–3,15,16^, the literature on this subject in the medical domain is relatively new and limited. Duckworth studied utilizing explainable boosting machines to characterize data drift for COVID-19 patients that were admitted to the emergency departments.^17^ By comparing the performance of the model on pre and post pandemic data, they were able to show a change in the relative importance of the features according to the Shapely Additive exPlanations (SHAP) values.^18^ They suggested that SHAP values can be used as a complimentary measure for data drift. Sharon Davis et al. proposed to use calibration drift as a measure of model deterioration over time and applied this method to data from department of Veterans Affairs hospitals using all-cause mortality as the endpoint.^19,20^ They analyzed the performance of multiple ML models and argued that, whereas the discremination power (in terms of the Area Under the Receiver Operating Curve) of all models remained relatively the same, the calibration power declined, as all models overpredicted the risks.^20,21^

In this study, we focus on assessing data drift in predicting sepsis, an important clinical application in which an ML-based tool is being used for sepsis prediction. We seek to define the characteristics of the data drift in sepsis prediction. To achieve this, we assembled a series of simulations capable of visualizing different forms of data drift in sepsis prediction models and measured the performance of two types of ML models trained to monitor patients for predicting the onset time of sepsis.

## 2. Materials and Methods

### 2.1. Dataset

We initially considered 113,037 retrospective patient encounters from a set of four hospitals in our network, from which we retained 112,972 encounters that occurred between January 2018 and January 2022. **Figure 1** shows the attrition chart and the number of encounters per site after exclusions. For ease of reference, the sites are numbered in a descending order based on the number of encounters at each site; hence site 1 has the largest number of encounters and site 4 has the smallest. **Supplementary Figure 1** shows the distribution of the number of encounters at all sites over the years that are considered in this study. The bulk of the encounters occurred between 2018 - 2021, and 2020 had the highest number of encounters.

**Figure 1.**
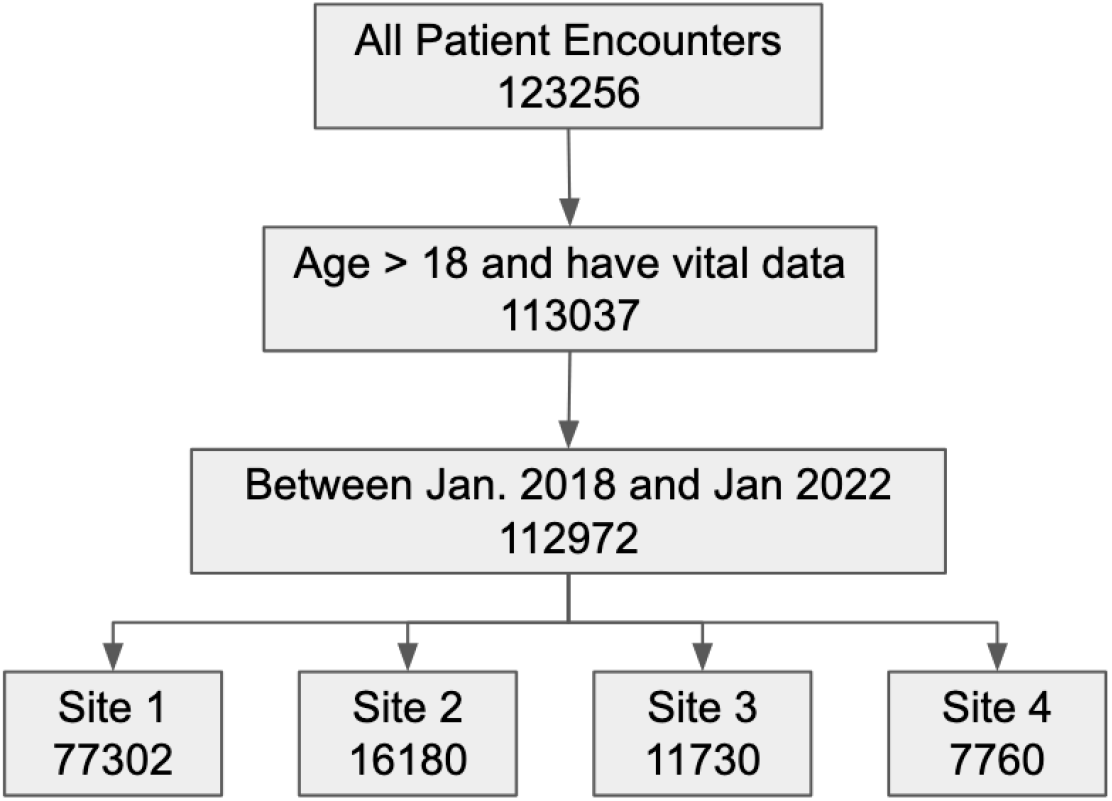
Data attrition chart.

### 2.2. Sepsis Definition (Target)

The definition of sepsis has undergone two revisions since 1991. The Sepsis-1 definition was primarily based on Systemic Inflammatory Response Syndrome (SIRS). This criteria was carried forward within the Sepsis-2 revision with additional criteria, including a requirement for the patient to have a confirmed or potential infection and additional clinical characteristics that resistant to fluid rescuicitation.^22^ Sepsis-3 considers Sequential Organ Failure Assessment (SOFA) as well as other clinical and laboratory observations.^5^ In this study, to induce a concept shift, we change sepsis definition from Sepsis-1 to Sepsis-3 to observe the effects of this change (together with the covariate shift) on the decay of model performance. It should also be noted that we were not able to fully adhere to the Sepsis-3 definition when trying to generate the labels, as we did not have the data required to calculate the Glasgow Coma Scale (GCS). However, for the purpose of the simulation, the labels are similar enough to the actual definition of Sepsis-3, for which the main manifestation is organ failure.

### 2.3. Covariates (Input Features)

With the exception of age and gender, all input features are time series data (**Table 1**). The recording of these features starts at the admission of the patient. Every recorded value has a timestamp that indicates when the value was taken. To effectively utilize the models to make predictions without too much missing data, we require that some of these features have at least one value during the time window in which the sepsis prediction is to be made. The required features are marked by asterisks in the table and include some of the patients’ most important and most frequently measured vital sign data: Systolic Arterial Blood Pressure (SysAPB), Diastolic Arterial Blood Pressure (DiasABP), Heart Rate (HR), Respiration Rate (RespRate), and Oxygen Saturation (SpO2).

**Table 1.** Input features to the ML algorithms. Five of the features, SysAPB, DiasABP, HR, RespRate, and SpO2, which are marked by asterisk, are required features, meaning that having at least one value of these features in the time series data is necessary to make a prediction.

|  | **Feature** | **Unit** |
| --- | --- | --- |
| 1 | Age | years |
| 2 | Gender (male, female) |  |
| 3 | Systolic Arterial Blood Pressure (SysAPB)* | mmHg |
| 4 | Diastolic Arterial Blood Pressure (DiasAPB)* | mmHg |
| 5 | Heart Rate (HR)* | BPM |
| 6 | Temperature (Temp) | °C |
| 7 | Respiration Rate (RespRate)* | BPM |
| 8 | Oxygen Saturation (SpO2)* | % |
| 9 | Partial Pressure of Oxygen (PaO2) | mmHg |
| 10 | Creatinine | mg/dL |
| 11 | Blood Urea Nitrogen (BUN) | mg/dL |
| 12 | Glucose | mg/dL |
| 13 | White blood Cell Count (WBC) | 1000/μL |
| 14 | Red Blood Cell Count (RBC) | million/mm<sup>3</sup> |
| 15 | Platelets | 1000/mm<sup>3</sup> |
| 16 | Neutrophils | % |
| 17 | Lymphocytes | % |
| 18 | Monocytes | % |
| 19 | Hematocrit | % |
| 20 | Lactate | mmol/L |
| 21 | pH |  |
| 22 | Aspartate Aminotransferase (AST) | U/L |
| 23 | Alanine Transaminase (ALT) | U/L |
| 24 | Bilirubin | mg/dL |
| 25 | International Normalized Ratio (INR) |  |

### 2.4. Machine Learning Models

Two MLAs are considered in this study, including an eXtreme Gradient Boosting (XGB) and a Recurrent Neural Network (RNN) with Gated Recurrent Units (GRU). RNN was selected over other neural network architectures due to the fact that the input features are time series data and RNNs are better suited for sequential data. Although RNN is more precisely referred to as a deep learning algorithm, here to ease the writing we refer to both models as MLAs. Both algorithms were included in this study to enable experimentation with different retraining methods. XGB cannot be incrementally retrained with the current implementation wherein the boosted trees are trained sequentially. As a result, an entirely new dataset needs to be built for XGB using old and new data. In contrast, RNNs, like other neural networks, can easily be retrained incrementally using only the new batches of data. This is similar to transfer learning techniques, in which a pre-trained model is further trained using new data.

### 2.5. Timely Sampling and Operation of the Models

The models trained for this study are not meant to simply be time agnostic risk classifiers. By time agnostic we mean predicting the risk of sepsis at “any time” in future [*t*, ∞) if *t* is the time at which the model makes a prediction. Instead, we are training our models to predict the probability of sepsis at a specific time in future (e.g., 24 hours from *t*). The actual time at which sepsis occurs in a patient is called the onset time; we denote this time as *T*_*s*_. For the purpose of these experiments, it can be assumed that *T*_*s*_ and all other times discussed herein are measured from the patient’s admission time. The choice of the time unit is arbitrary, and all of the times are measured in hours. *W*_*p*_ will denote a fixed amount of time before *T*_*s*_ at which we wish to predict if sepsis happens at *T*_*s*_; that is, the goal is to predict sepsis at most at *T*_*s*_ - *W*_*p*_. Of course, during the training, we know the actual *T*_*s*_ and build our training samples with respect to it, but in deployment, *T*_*s*_ is unknown and at each time *t* we make a prediction about the probability of sepsis at *t + W*_*p*_, with the goal of making an accurate prediction for sepsis onset. In this way, we provide healthcare practitioners a window of *W*_*p*_ hours to intervene and prevent sepsis from happening if a patient is predicted to be highly at risk.

In this setting, the model continuously runs and monitors patients as their data stream is generated. The model starts running after a patient is admitted to the hospital and as soon as at least one measurement for each of the required features is available. It continues to predict the probability of sepsis at every time *t* when a new value is available for any of the features. The model continues to operate until monitoring of the patient is stopped by the practitioners, due to discharge, mortality, or another reason. The sepsis probability between the two consecutive measurements of any of the features is assumed to be the same, as we do not have information to deduce otherwise if none of the features have changed since the last time.

The time series data continues to be recorded after a patient’s admission, and the length of the data for each patient continues to grow until discharge or mortality or discontinuation of recording due to any other reason. The next question we seek to address is the manner in which this input data is fed to the models to obtain a prediction. The large variation in the length of the time series features for each patient and the older parts of the time series data that have likely less impact on the patient’s present condition makes it a challenge to develop the ML models. As such, we utilize a look back window to extract input features for the model and denote this window by *W*_*l*_. It is defined as the time window before the present time (at which the model makes prediction) where the data is relevant. This means that at each time *t* when the model runs, it uses data from the interval [*t* - *W*_*l*_, *t*] to make a prediction if sepsis occurs at *t + W*_*p*_. In this research, we set *W*_*l*_ = 48 hours and *W*_*p*_ = 24 hours based on feedback from the hospitals in our network and also our experience with analysis of the data.

Using the above windows, the training samples are constructed. Every patient encounter potentially has many time steps in the time series data, meaning that several samples can be taken from every patient encounter. To build the samples, we use a sliding window approach that iterates at every time step *t* in the patient’s data and builds a sample that has features taken from [*t* - *W*_*l*_, *t*]. This window labels the data according to whether the patient is positive or negative at *t + W*_*p*_. The samples that are built for the early time steps right after admission are backfilled if there is an inadequate amount of data. Additionally, because the intervals between every two consecutive updates of any of the features varies, the values in the sampling window are binned at every hour in the window to make a fixed size input of the size *W*_*l*_ for each feature, that is the time series feature matrix for a single sample has the the dimensions *n*_*f*_ X *W*_*l*_ where *n*_*f*_ is the number of input features (except for age and gender) in **Table 1**.

The total number of samples *m* that are obtained in this way is several times bigger than the total number of encounters in **Figure 1**, as each patient contributes to more than one sample. A negative patient will always produce negative samples since the patient never becomes positive. A positive patient can produce both positive and negative samples depending on the time *t* at which the sample is made. To prevent the data leak, the samples for positive patients are taken only from the data prior to the actual sepsis time *T*_*s*_ (minus the prediction window *W*_*p*_). We exclude patients who were positive in the first 24 hours after admission. To maintain the same rate of positive patients in the samples as the encounters, we limit the number of negative samples that are taken per patient. Finally, to reduce the bias that might be caused by patients who have a lot of data (e.g., very long duration of stay), we limit the total sampling window per patient to the first month since admission.

In the case of XGB, the *n*_*f*_ X *W*_*l*_ time series sample is converted to a set of summary statistics features and concatenated to the demographics features (age, gender) to form the input vector to the model. The summary statistics features include the minimum, mean, maximum values of the feature, as well as recent values in *W*_*l*_. The RNN model on the other hand uses the time series sample *n*_*f*_ X *W*_*l*_ as is. It has two layers of GRU, with *W*_*l*_ units in each layer for each time step in the input. The two GRU layers are then followed by a fully connected (dense) layer. The demographics inputs in this case are directly fed to the fully connected layer together with the outputs of the last GRU layer. The RNN models were regularized using drop out.

### 2.6. Measuring the Performance

Predicting the onset time of a risk (specific time in future) is more challenging than the time agnostic risk classification (any time in future). In such cases, the time agnostic definitions of true positive (TP), false positive (FP), true negative (TN) and false negative (FN) could be misleading and provide optimistic results compared to reality. In this research we use a timely definition of these concepts. To simplify the presentation, let’s call every *positive prediction* done by the model an *alert*. **Table 2** shows the time agnostic versus timely definitions of positives and negatives. As can be seen, only the positive patients who were alerted before the onset time are counted as TP. A late alert for a positive patient is counted towards FN, because it is too late. Subsequently, the definitions of sensitivity, specificity, and positive predictive value (PPV) are based on the timely measurement of TP, FP, TN and FN.

**Table 2.** Comparison of time agnostic versus timely definition of TP, FP, TN and FN. In the timely definitions, the predictions (alerts) are counted with regard to the ground truth onset time.

| Time agnostic | Timely |
| --- | --- |
| **TP:** Septic patients who got an alert | **TP:** Septic patients who got an alert before the true onset time. |
| **FP:** Non septic patients who got an alert | **FP:** Non septic patients who got an alert |
| **TN:** Non Septic Patients who never got an alert | **TN:** Non septic patients who never got an alert |
| **FN:** Septic patients who did not get any alerts | **FN:** Septic patients who did not get an alert before the true onset time, i.e., septic patients who never got an alert + septic patients who got an alert after the onset time |

### 2.7. Simulation Methodology

To simulate the effects of data drift on the performance of the models, the following scenarios are considered.

- When there is a sudden and potentially catastrophic major event (e.g., COVID-19)
- When there is no apparent concept shift (labels are the same), but there could be a covariate shift
- When there is a concept shift (change of labels)

In executing all of the above scenarios, we observe the changes in the prevalence of septic patients (prior probability shift), the Area Under the Receiver Operating Characteristic curve (AUROC), and the average calibration value of the models. We also observe the performance metrics for fixed operating points such as the normalized ratio of positive predictive value (PPV) at a fixed sensitivity = 0.8 for all models. Because PPV changes directly with prevalence, we chose a normalized measure of PPV, that is PPV divided by prevalence, called Lift,^23^ as a more desirable metric.

In order to perform simulations, we sort the encounters by admission time and split the encounters at a specific time (split time = *T*_*split*_). We retain the encounters that occur prior to the split time (pre-split encounters *E*_*pre*_) for the baseline models. Those that occur after *T*_*split*_ (post-split encounters *E*_*post*_) are used for retraining simulation and measuring the data drift. The training and validation datasets for baseline models are randomly created from *E*_*pre*_ by 80:20 split. For the retraining phase, *E*_*post*_ is first divided into equal partitions *E*_*i*_ with an identical number of patient encounters. These partitions are then sequentially used to simulate the retraining phase. At every simulation iteration *i*, the encounters *E*_*i*_ are used to retrain or tune the models depending on the model type. The encounters *E*_*i+*1_ are used for testing the performance of the baseline and retrained models at this iteration to demonstrate data drift. This process is illustrated in **Figure 2**.

**Figure 2.**
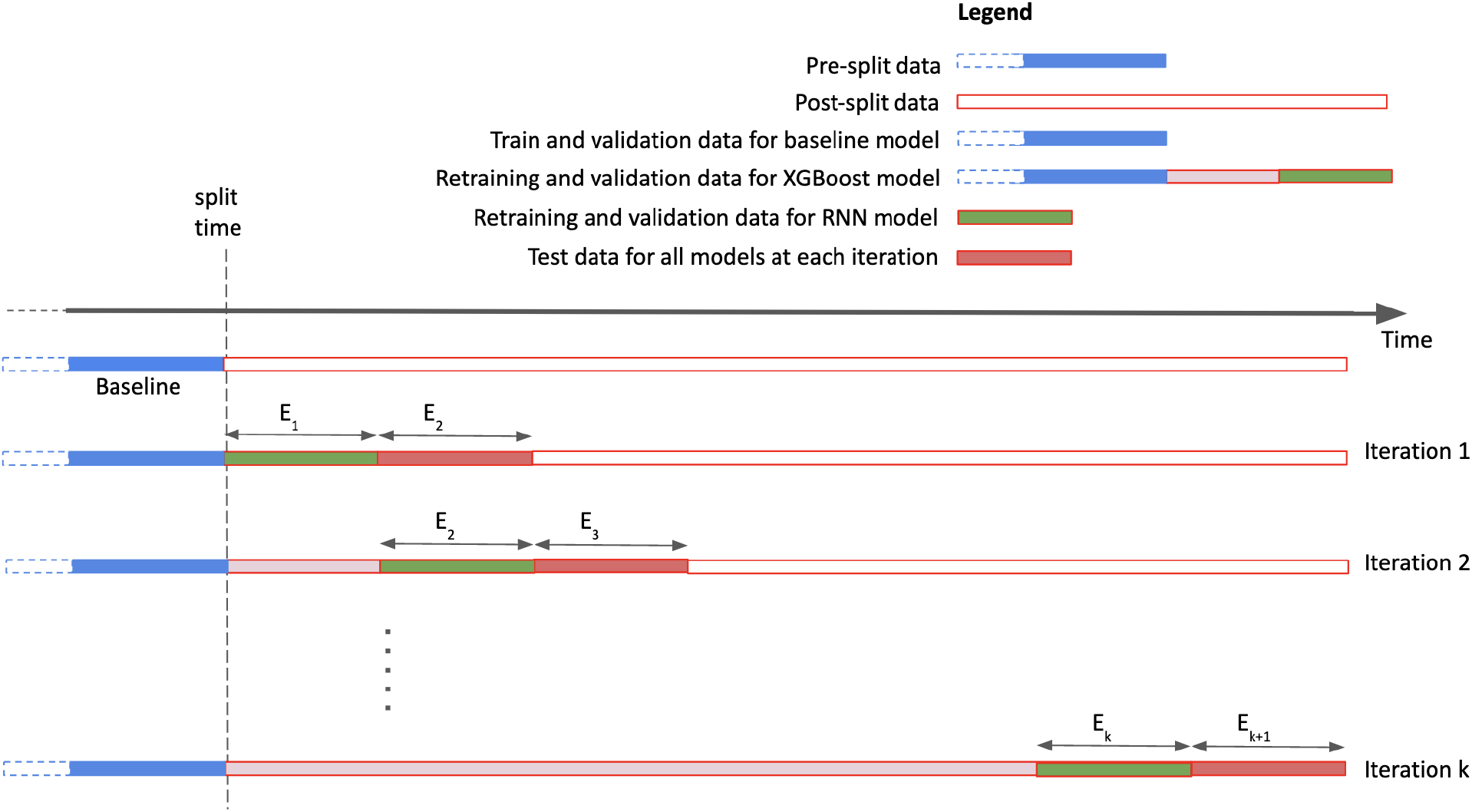
Data splits for the simulation study. The lengths of the bars are not to scale and are for illustration purposes only.

In the case of XGB, a baseline model is trained on pre-split data and then a set of retrained models are trained on the post-split data. During retraining, the initial pre-split encounter set is continuously augmented with new data and at each simulation step a new model is fully trained on the concatenation of the old and new data. We also explored an alternative method of retraining by using a sliding window in which the oldest encounters are dropped as the new ones are added, however, we do not report it herein as it did not make any notable difference in our simulations.

In the case of RNN, a baseline model is trained on the pre-split data which is cloned before the retraining starts. The cloned model is then continuously retrained using post-split data. Only each individual partition is used for retraining at each step. The RNNs can be tuned in two different ways. The first resembles transfer learning, in which we freeze the first layers of the model and only allow the final layers to be trained. The second is to allow the entire model to be trained with a very small learning rate. The former method showed better performance and thus, the results that are reported in this paper are based on this method.

Using these models, we compared the performances of retrained models against the baseline models under data drift. In all models built in this study, we used regularization, early stopping, and other hyper parameter tuning methods to train the best possible model.

As a final note, we intentionally used the term “encounter” in naming pre-split *E*_*pre*_ and post-split *E*_*post*_ data, to indicate that the splitting is done based on the encounters, not the input samples for the models, to be consistent at the encounter level. Once the encounters are split, the input samples are made based on the methodology of section 2.5. Also, since the pre-split data is used for the baseline models and the post-split data is used for the retrained models, we will use the terms pre-split and baseline as well as post-split and retrained (or simulated) interchangeably in the rest of the paper.

## 3. Results

### 3.1. Major Event

For this simulation, the label definition for the baseline and the retrained models remains the same and uses the Sepsis-3 label. **Figure 3** shows the number of daily new COVID-19 cases in the United States since the pandemic began in 2020. Although the pandemic was declared a public health emergency in the United States in March 2020, we use July 2020 as the split time for the major event in our simulation. We selected this split time to ensure that we had adequate patient data for our simulations, which would not have been possible at the start of the pandemic. The use of March 2020 as the split time would have significantly reduced the baseline training data in our dataset. Additionally, July 2020 is the point at which COVID-19 case counts in the United States exceeded 3 Million.^24^ As such, the July 2020 wave of the virus yielded significantly more cases and hospitalizations and the strain on the healthcare system and the potential for this strain to influence data drift can be seen more clearly from July 2020-on.

**Figure 3.**
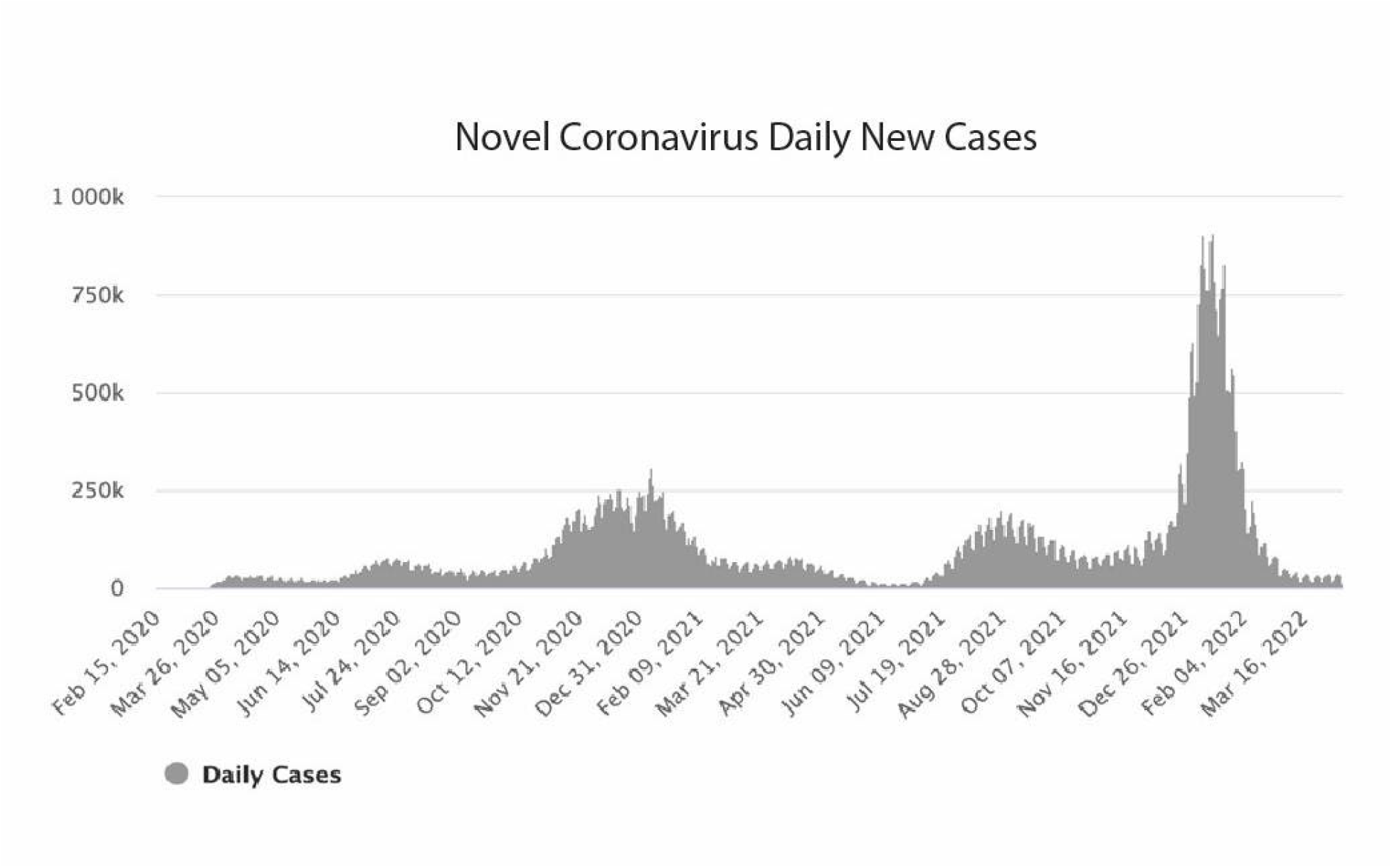
The number of daily new cases in the United States.^25^

The split results in |*E*_*pre*_ | = 49,409 encounters for the baseline models and |*E*_*post*_ | = 63,563 for the retraining simulation. Each retraining iteration uses data from |*E*_*i*_| = 6000 newly encountered patients. This results in 10 simulation steps, but since only a fraction of the 6000 encounters will be available for testing the last step, we discard the results of the 10th iteration to consistently have the same number of encounters for all test sets *E*_*i+*1_. Choosing 6000 encounters for each iteration was done arbitrarily to have about two months of intervals between iterations. With this way of splitting, we get |*XY*_*pre*_| = 106,759 and |*XY*_*post*_| = 238,203 samples for the XGB simulation and |*XY*_*pre*_| = 170,791 and |*XY*_*post*_| = 370,686 samples for the RNN simulation. We have used the symbol *XY* to denote the samples that constitute inputs and targets to the machine learning models.

As seen in **Figure 4A**, the baseline XGB model outperforms the RNN model specially for operating points that are near the high specificity region of the receiving operating characteristic (ROC) curve. This generally turns out to be the case throughout the rest of the simulations presented in this paper, where in most cases the XGB model performs either better or as good as the RNN model, which may be due to the use of engineered features (summary features) in the XGB samples that results in better predictive power compared to the actual time series features that are used in the RNN samples. It should be emphasized however, that the reason we have used two types of models in this paper is not to find the most optimal model architecture, but instead to have two different architectures that require different methods of retraining, to see if it may have an impact on compensating for the data drift.

**Figure 4.**
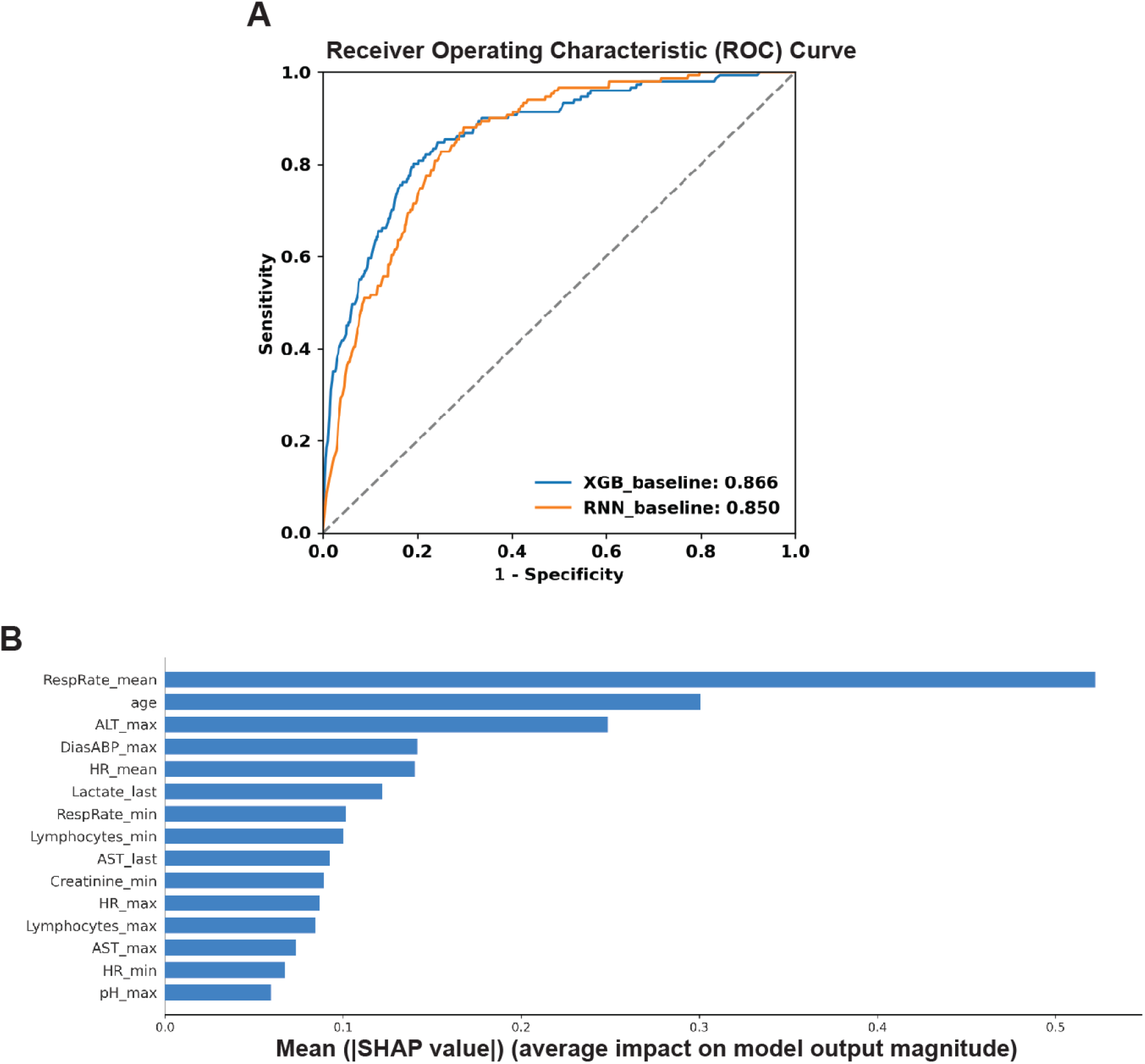
Major event simulation **A**. Timely ROC curves for baseline XGB and RNN models using data from 3000 randomly chosen patients from the baseline data. **B**. Most important features according to SHAP values of the baseline XGB model.

The SHAP plot ^18,26^ that represents the most important features of the XGB model is shown in **Figure 4B**. Here, and in the rest of the paper, we only report the distributions of the features across the baseline and retraining datasets (pre- and post-split) only for the most important features according to the baseline XGB models’ SHAP plots. **Figure 4B** shows that the engineered features from RespRate, age, ALT, DiasABP, HR, Lactate, Lymphocyte, Creatinine, AST and pH are among the most important features of the model. The names of the features in the SHAP plot are suffixed by the statistical method that was applied to obtain that particular engineered feature, for example min for minimum, max for maximum, and last for the last value in the time series window. The **Supplementary Table 1** shows the distribution of the features in each simulation iteration for all encounters and all time-series values for that feature in that iteration.

The rates of prevalence (rate of positive patients) per each iteration are shown in **Figure 5A**. The average baseline prevalence is also shown for reference. It demonstrates a significant gap between the average baseline prevalence and the iteration prevalence. This signals a prior probability shift (change of P(Y)) in the dataset which might be due to COVID-19 as physiological responses to Sepsis and Acute Respiratory Distress Syndrome (ARDS) share some similarities, such as leukocyte activation.^27,28^ **Figure 5B** Shows the changes of AUROC per iteration for retrained XGB models. The overall average of the AUROC for all iterations is also shown in the figure’s legend. The error bars represent the 95% confidence intervals, which we have included in all subsequent figures. **Figure 5C** shows the changes of the average model calibration for the XGB models. The calibration value at each iteration is the average Observed to Expected ratio (O:E) of risk (being positive). ^29^ **Figure 5D** shows the changes of Lift at a single operating point with sensitivity = 0.8 per iteration for the baseline and retrained XGB models.

**Figure 5.**
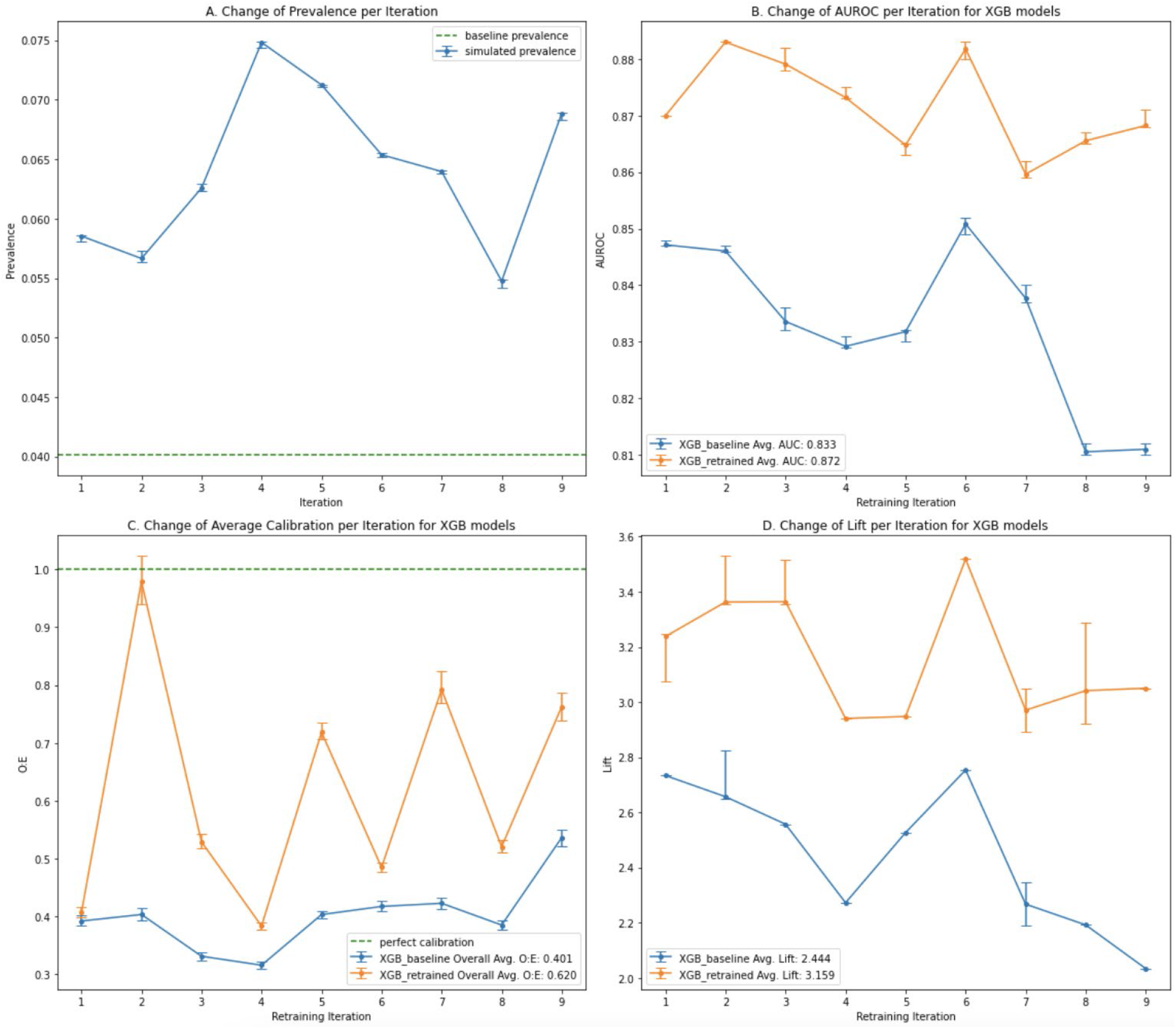
Major event simulation. **A**. Change of prevalence for each iteration in the post-split data. The dashed line is the average baseline prevalence. **B**. Changes of AUROC per iteration for XGB models. Error Bar: 95% CI. The values at the end of the period (iteration 9) are 0.811 for the baseline and 0.868 for the retrained XGB model (reported in the abstract). **C**. Changes to the average calibration per iteration for XGB models. The overall O:E ratio for all iterations is shown in the legend. **D**. Changes of Lift at sensitivity = 0.8 per iteration for XGB models.

Evidently, **Figure 5** shows a marked difference between the baseline and the retrained models. This is likely attributed to an overall shift in the data. While the prior shift is very significant for the most part, the distribution of the covariates, with the exception of a few cases, does not show dramatic changes as seen in the **Supplementary Table 1**. This is a further indication that isolated comparison of the distribution of covariates may not be sufficient, as it does not reflect the overall shifts in the joint distribution of the data. As such, performance metrics such as AUROC and calibration are more useful to highlight the data drift. It can also be seen that the retrained models perform significantly better than the baseline model for all iteration steps, and the performance gap tends to increase per iteration. These results highly demonstrate the necessity for a major retraining of the models in case there is a big event such as COVID-19 pandemic.

The **Supplementary Figure 2** shows the AUROC and calibration plots for the RNN models. Note that the prevalence plot does not depend on the model and will be the same, and we skipped the Lift curve to reduce the number of plots in the supplementary material. The AUROC plot for the RNN models shows mixed results, although on average the retrained models seem to be doing better than the baseline model. For the RNN models, the calibration curve of the retrained model is significantly better than the baseline model at all points, even though the calibration values for both RNN models are poor compared to the XGB models. The low calibration of the RNN models is primarily due to lower predicted probability values in these models.

### 3.2. Covariate Shift

Even without any major unforeseeable event, model performance may still degrade because of covariate shift. To measure the effects of covariate shift, we produce the same set of plots as in the previous section to evaluate for whether a covariate shift occurs in the data and the resulting impact on model performance. The split time used in this and subsequent simulations is 1^st^ January 2021. This time was chosen arbitrarily to provide us with approximately one year of data for the simulation part.

The split results in |*E*_*pre*_ |= 74,426 encounters for the baseline models and |*E*_*post*_ | = 38,546 for the retraining simulation. Each retraining iteration uses data from |*E*_*i*_| = 4000 newly encountered patients, which is less than the previous simulation as we have less data for simulation in this case and wanted to generate enough points on each curve. As in the previous simulation, we discarded the result of the last iteration to consistently have the same number of encounters for all test sets *E*_*i+*1_. With this way of splitting, we get |*XY*_*pre*_| = 202,880 and |*XY*_*post*_| = 143,382 samples for the XGB simulation and |*XY*_*pre*_| = 320,517 and |*XY*_*post*_| = 222,989 samples for the RNN simulation.

The performance and most important features of the baseline XGB model for this scenario are shown in **Figure 6**. The distribution of the most important features in this case is shown in the **Supplementary Table 2**. An identical category of plots to those used in the previous simulation for this scenario are shown in **Figure 7**. The prevalence remains substantially higher than the baseline and the performance gap between AUROC of the baseline and the retrained models is still maintained, though the gap is slightly smaller than the previous simulation. The retrained models also generally show better calibration than the baseline model. These facts suggest that covariate shift can occur in sepsis and similar clinical settings, and the shift demonstrates more of a fluctuating nature than a continuously evolving one. This is in contrast to what might be seen in models trained for spam detection, for example, where the model has to cope with the shifting strategies of an intelligent adversary. ^30^ These shifts are better represented in the model performance plots than the marginal distributions of each individual feature (**Supplementary Table 2)**. Performance gaps at a single operating point (Lift) are not as widely separated as in the previous simulation. This, once again, highlights that monitoring model performance requires a multi-faceted approach. The AUROC and Calibration plots for the RNN models are shown in **Supplementary Figure 3** where we see mixed results. The retained RNN models show better calibration but worse AUROC where we kept the model architecture fixed.

**Figure 6.**
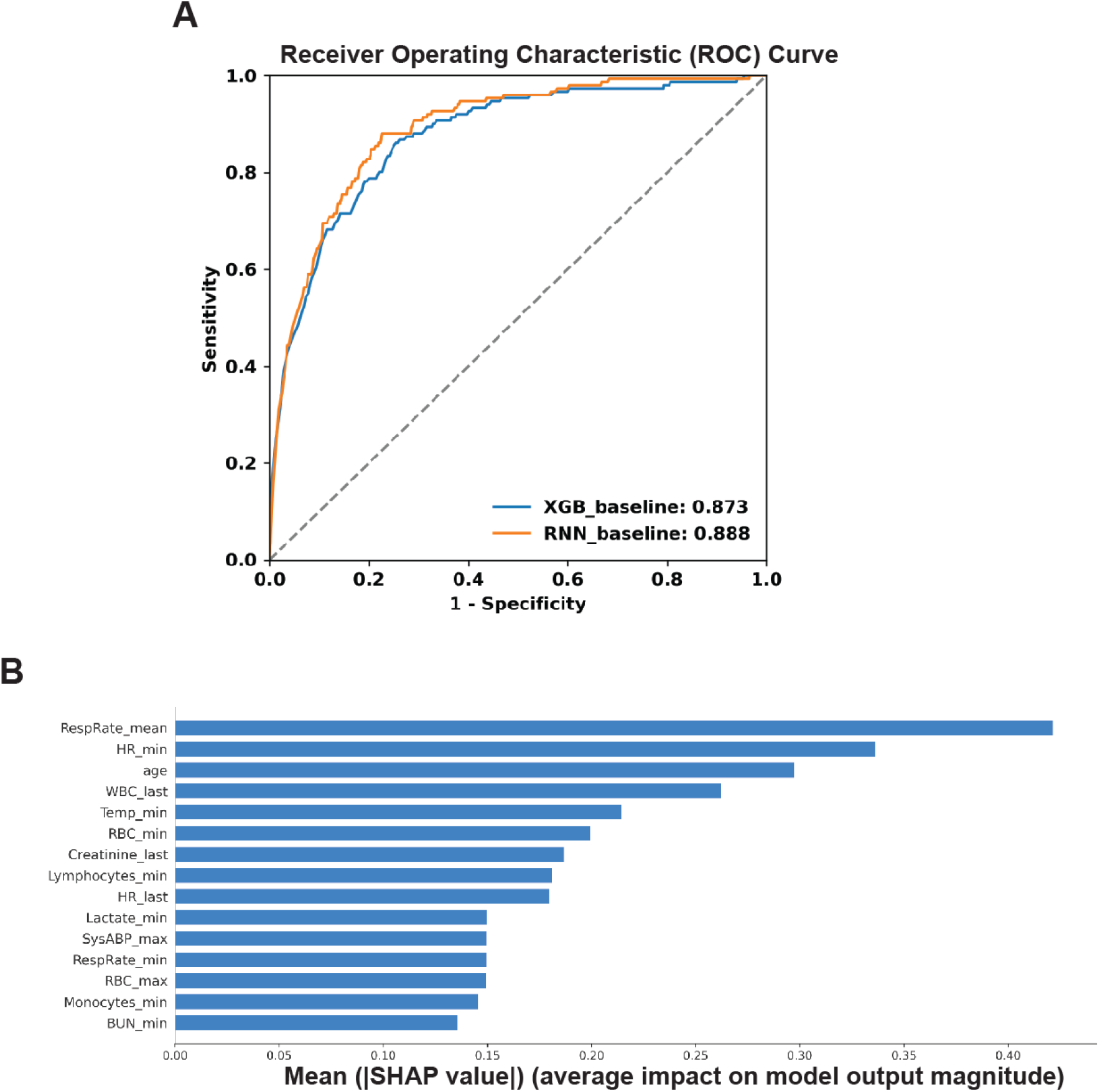
Covariate shift simulation. **A**. Timely ROC curves for baseline XGB and RNN models using data from 3000 randomly chosen from the baseline data. **B**. Most important features according to SHAP values of the baseline XGB model.

**Figure 7.**
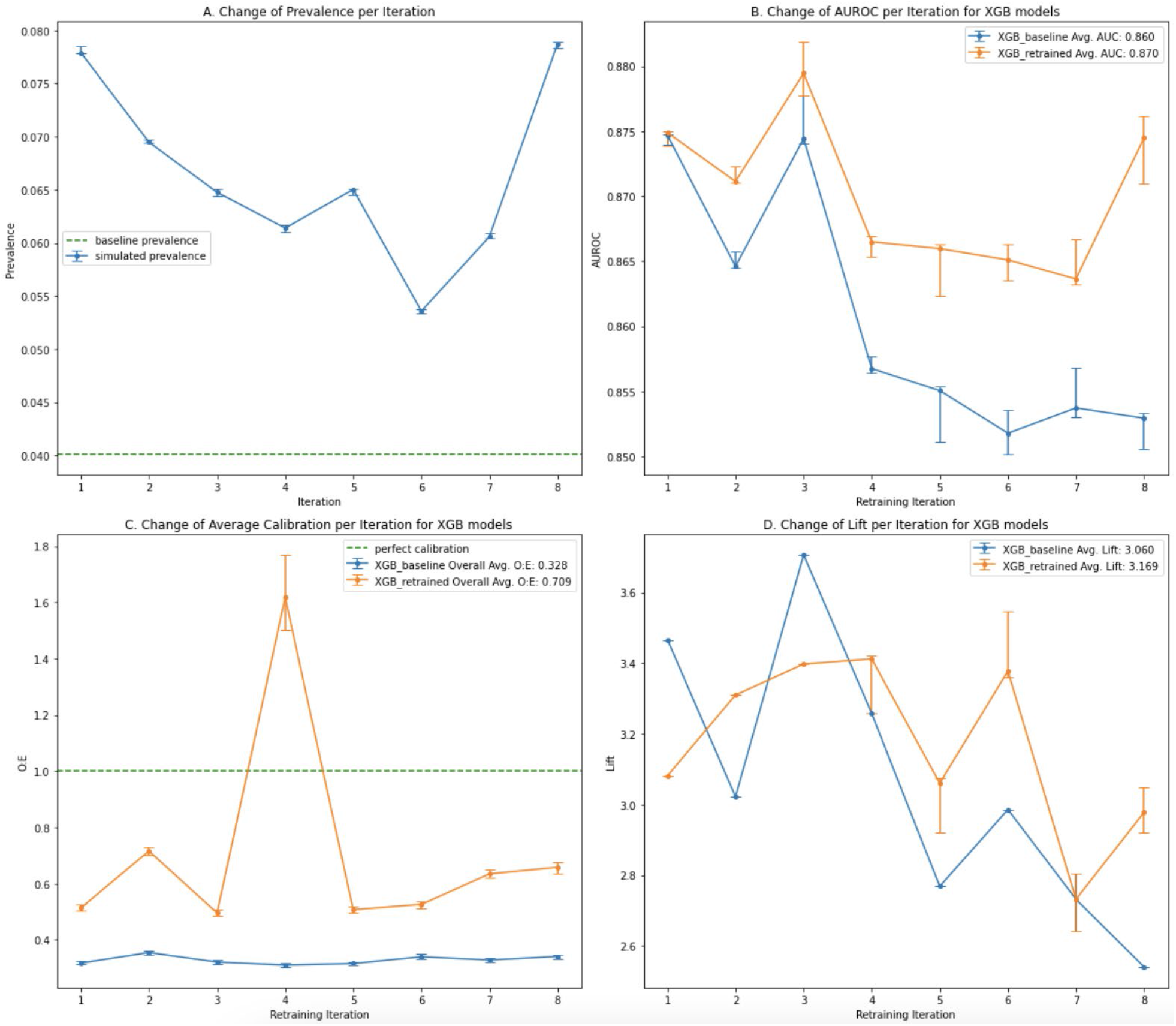
Covariate shift simulation. **A**. Change of prevalence for each iteration in the post-split data. The dashed line is the average baseline prevalence. **B**. Changes of AUROC per iteration for XGB models. Error Bar: 95% CI. The values at the end of the period (iteration 8) are 0.853 for the baseline and 0.874 for the retrained XGB model (reported in the abstract). **C**. Changes to the average calibration per iteration for XGB models. The overall O:E ratio for all iterations is shown in the legend. **D**. Changes of Lift at sensitivity = 0.8 per iteration for XGB models.

### 3.3. Concept Shift

The split time and the number of patient encounters in this simulation are identical to those described in section 3.2., i.e., January 2021, |*E*_*pre*_ |= 74,426 and |*E*_*post*_ | = 38,546. The number of post-split samples is also the same because we use the same label, sepsis 3, for the post-split part. The number of samples for the baseline models however are slightly different since we use a different label, sepsis 1, for the baseline models which triggers slightly different criteria to filter samples and results in |*XY*_*pre*_| = 224,084 for the XGB and |*XY*_*pre*_| = 335,197 for the RNN baseline models.

In this simulation, we attempt to induce a concept shift by training the baseline models on sepsis 1 label and then retraining and simulating the performance on sepsis 3. However, inducing the concept drift in this way poses some unique challenges.

First, it is not possible to fully isolate the effects of covariate shift from concept shift, as covariate shift in the data cannot be fixed. As a result, what we observe here is the combined effect of the covariate shift and the concept shift. By using the same split time, the distribution of the covariates will be essentially the same as the **Supplementary Table 2**, and we skip reproducing it and the SHAP plot for this scenario, although the list of the most important features may be different because a different sepsis label is used for the baseline model. The prevalence change for the post-split data is also identical to that depicted in **Figure 7.A**, but the baseline prevalence is different due to a different sepsis label for the baseline data.

Additionally, concept shift in practice may be a gradual process, which we cannot replicate here. By changing the label from sepsis 1 to sepsis 3, on post-split data, we are inducing a sudden concept shift in the data. However, this simulation can still be useful because in the case of sepsis prediction, a gradual concept shift is unlikely to occur in real life scenarios, as this would require the perception of sepsis to continually change over time. It is more likely that a new consensus on the definition of sepsis would evolve over several years, as with previous variations to the definition of sepsis, which will then be adopted in practice.

The simulation results for the XGB models in the concept shift scenario are shown in **Figure 8**. As before, we gradually augment the samples for the XGB models with the new data, but this will have some ramifications in this case. The issue is that the baseline models are trained on old labels, but the new batches of data carry the new label. As such, we see mixed results for AUROC as shown in **Figure 8.B**. The AUROC of the retrained models, particularly for the first few iterations, is actually lower than the baseline model due to this mixing of labels. The similarity between the two labels allows the baseline models to maintain a reasonable level of performance for a while. The retraining performance appears to catch up towards the end of the iterations, but we do not have enough data to continue to observe this trend. The sliding window method (dropping the old encounters as the new ones are added) also did not improve the results and will not be reported here. Plots for the RNN models are shown in **Supplementary Figure 4** and demonstrate similar trends.

**Figure 8.**
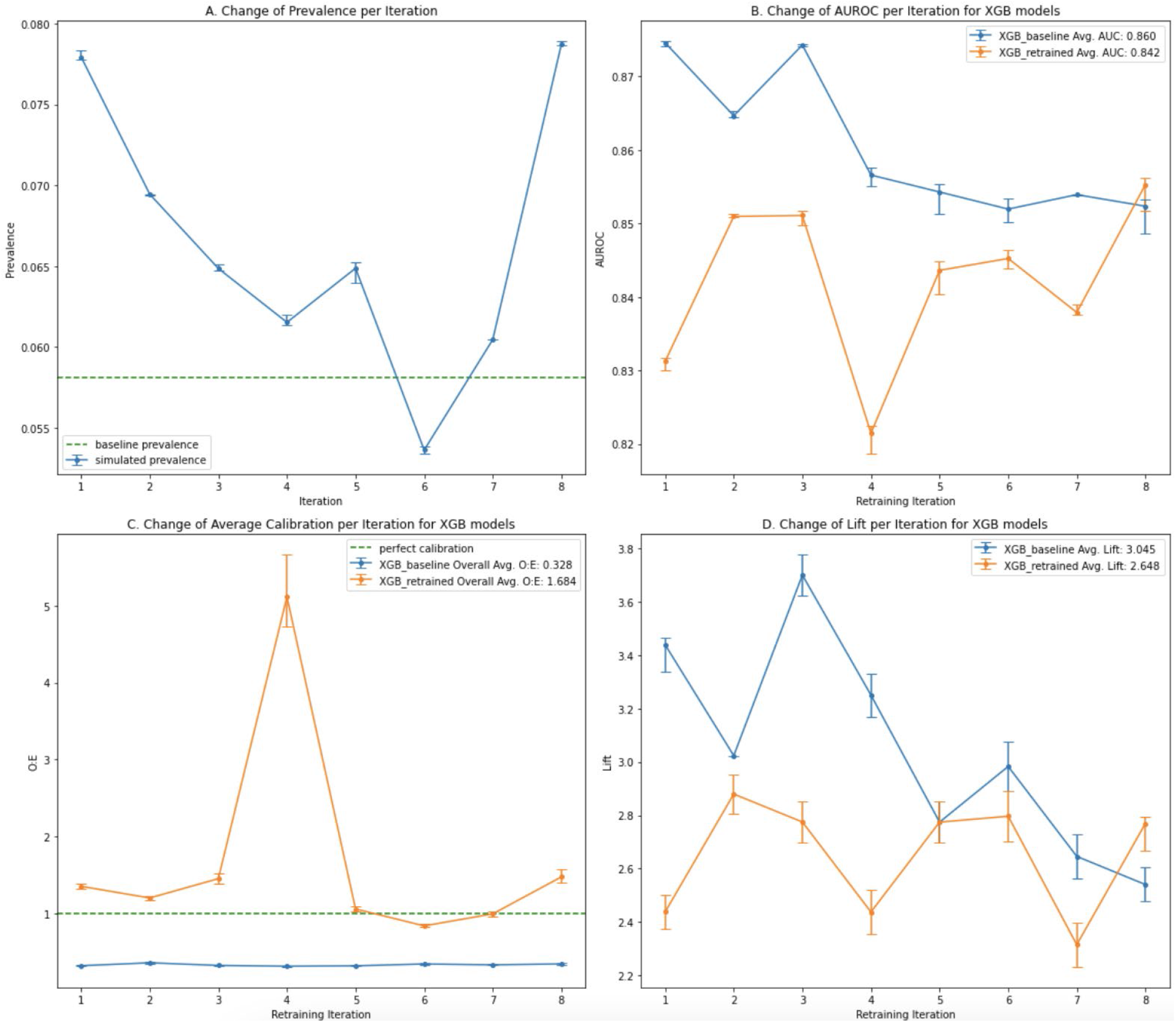
Concept shift simulation, mixed labels. **A**. Change of prevalence for each iteration in the post-split data. The dashed line is the average baseline prevalence. **B**. Changes of AUROC per iteration for XGB models. Error Bar: 95% CI. **C**. Changes to the average calibration per iteration for XGB models. The overall O:E ratio for all iterations is shown in the legend. **D**. Changes of Lift at sensitivity = 0.8 per iteration for XGB models.

A better approach for retraining in this situation may be to fully relabel the dataset with the new labels and retrain the model. In this way, we do not incrementally add data with new labels to the dataset that has old labels; rather, we rebuild the entire training dataset with the new label. As this method does not involve any incremental retraining, we focus on the XGB model type as it is the model type that requires a whole dataset in retraining, and its performance has been as good as RNN in all previous simulations. A priori, we can hypothesize that the retrained models using fully relabeled data should perform better in this way, and we demonstrate this in **Figure 9**, where the AUROC and the calibration curves of these fully relabeled and retrained XGB models are significantly better than the baseline model. The lift curves, however, are not very distinctive.

**Figure 9.**
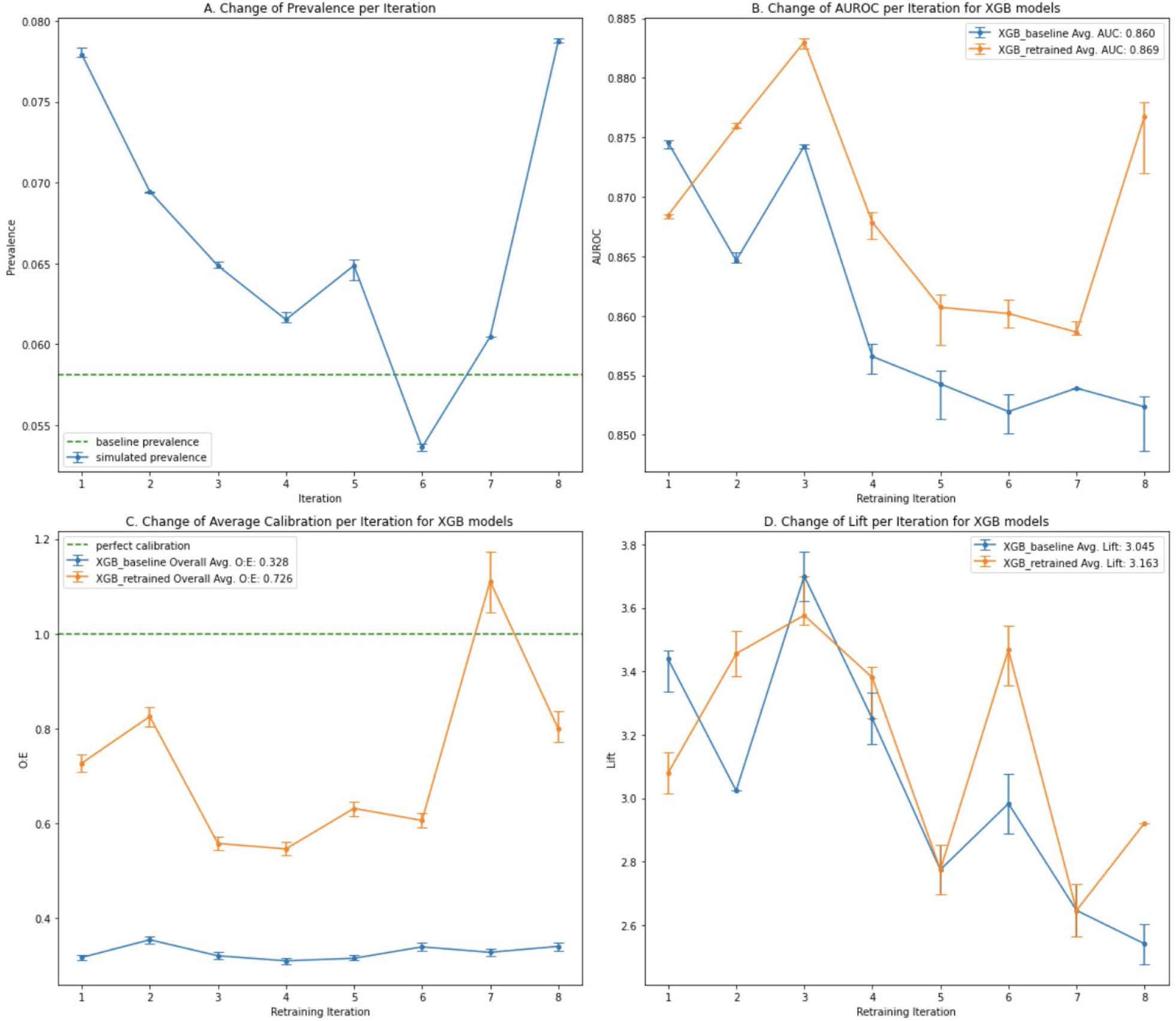
Concept shift simulation, full relabeling. **A**. Change of prevalence for each iteration in the post-split data. The dashed line is the average baseline prevalence. **B**. Changes of AUROC per iteration for XGB models. Error Bar: 95% CI. The values at the end of the period (iteration 8) are 0.852 for the baseline and 0.877 for the retrained XGB model (reported in the abstract). **C**. Changes to the average calibration per iteration for XGB models. The overall O:E ratio for all iterations is shown in the legend. **D**. Changes of Lift at sensitivity = 0.8 per iteration for XGB models.

## 4. Discussion

The results of our simulations reveal that data drift is indeed present in a clinical condition such as sepsis and that retraining is required to maintain good model performance. Whereas the distribution of data in sepsis prediction does seem to change and sometimes be even exacerbated by major events such a COVID-19 pandemic, it does not seem to happen on a continual and gradual basis.

Although a major covariate shift is seen during a major event, the shift is somewhat less pronounced if the baseline model is not fully trained on the data prior to the major event. This is observed at least for the duration of the simulations we used here. More substantial covariate shifts may be manifested however if one could run a longer simulation in future research. Prior probability shift seems to be common and fluctuates significantly by time, particularly if the duration contains a pandemic. Continuous and gradual shift in concept for a disease like sepsis would not normally occur, as its definition should not change frequently. A change of definition, or specific interpretations of the standard definitions by healthcare provides, will be a basis for concept shift. This would likely require a full relabeling and retraining rather than incremental fine tuning and adaptation. Although the relabeling of the entire dataset is relatively easy in the case we have presented in this paper, this could be a major limitation for other applications due to the cost and time required for relabeling, for example in cardiovascular imaging.^31^ Handling the applications in which relabeling is a big challenge could be an avenue for future research.

The simulation curves presented in this paper demonstrated that a single performance monitoring curve may not be adequate to measure model performance, as different curves may reveal different aspects of model performance. With enough computing resources, one can also visualize the shifts in feature importance (based on SHAP values) as an added measure. Average calibration values that we presented in this paper are known as weak calibration values. In future work, stronger forms of calibration curves that are more computationally demanding, such as estimated calibration index, ^29^ can be explored.

The method of retraining of the RNN model, which only focuses on freezing some of the layers and tuning the learning rate may be inadequate, as evidenced by poorer performance of the retrained models compared to the baseline model in many cases. However, the substantial efforts that are required to re-adjust the RNN architecture at each iteration step made it impractical to do it in this research solely for the purpose of obtaining a better retrained model, particularly in light of having better performing XGB models. In this research we focus only on retraining methodology with respect to the data utilization to demonstrate such effects.

Our first set of limitations in performing these simulations stems from the dataset itself. We have missing features in the dataset, some of which affect the sepsis definition. Future work should use more data for the baseline (pre-split) and the simulation (post-split). This will allow running a full simulation that is entirely in either pre-pandemic or post-pandemic data. The lack of data may also be problematic for drawing definitive conclusions on the length of time it takes for the data drift to be noticeable in sepsis prediction, and accordingly, to build a model monitoring and retraining schedule that is best adapted to it. The pre-pandemic dataset for the major event simulation may also be inadequate and adversely affect the baseline performance.

For future research, we intend to verify our simulations with actual randomized trials that will be carried out in the four hospitals whose data was used in these simulations. This will be done via a two-arm experiment that will compare the performance of the baseline model with the retrained models. Another interesting future research question that may be answered by simulations, given that there is a dataset with enough features, is to be able to find out if the peaks and dips in the prevalence and performance of the models are associated with the Delta and Omicron variants of COVID-19.

### 5. Conclusion

In this paper, we presented a series of simulations intended to demonstrate the nature and dimensions of data drift in sepsis prediction. Our simulations revealed several characteristics of data drift in sepsis prediction and the impacts of the retraining methods on model performance under such circumstances. These findings may inform future research on the development and refinement of better patient monitoring systems to predict disease states in dynamic scenarios, which can improve both the cost and quality of care.

## Data Availability

This study used data from a proprietary commercial database and thus, cannot be shared

## Summary Table

| What was already known on the topic | What this study added to our knowledge |
| --- | --- |
| • Data drift can affect the performance of machine learning algorithms.<br>• Machine learning models have been deployed in clinical settings to predict the onset of disease conditions.<br>• There is a dearth in research on the impact of data drift on clinical prediction models that use machine learning for making predictions. | • We simulated three scenarios for data drift – covariate shift, concept shift and the occurrence of a major event – and studied their impact on ML-based clinical sepsis prediction models<br>• Significant data drift is present in sepsis data, and this can have an impact on model performance<br>• Retraining methodology in terms of how the new data is consumed and labeled is important for maintaining good model performance<br>• Retrained XGB models that used engineered features and an easily modifiable architecture performed better than the RNN models that used time series features and a fixed architecture |

## Acknowledgements

We would like to thank S. Shokouhi for reviewing and editing the manuscript.

