## Supplemental Material for "Assessing the effects of data drift on the performance of machine learning models used in clinical sepsis prediction"

### Appendix: Supplementary Materials

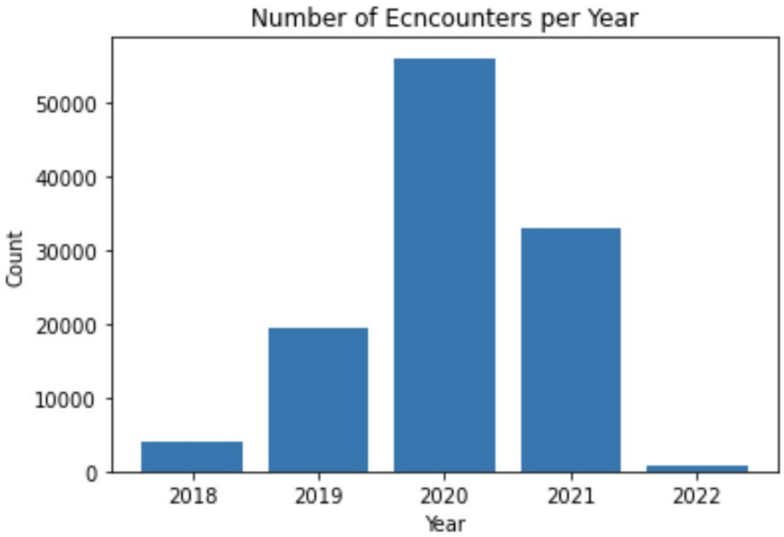


**Supplementary Figure 1.** Distribution of number of patient encounters per year of study, for all sites.

**Supplementary Table 1.** Major event simulation. Distribution of the most important features. The values are in the format of mean (std). Iteration 0 is the baseline distribution.

| Iteration | RespRate (BPM) | ALT (U/L) | Diastolic BP (mmHg) | Lactate (mmol/L) | Lymphocytes (%) | AST (U/L) | Creatinine (mg/dL) | pH | Age (years) |
|---|---|---|---|---|---|---|---|---|---|
| **0** | 17.73 (4.78) | 45.97 (59.31) | 69.93 (14.28) | 2.30 (2.03) | 18.73 (11.57) | 46.85 (55.13) | 1.37 (1.98) | 7.38 (0.11) | 50.42 (19.81) |
| **1** | 17.81 (4.50) | 46.80 (59.93) | 70.71 (13.86) | 2.32 (2.17) | 18.62 (11.13) | 42.09 (54.17) | 1.33 (1.37) | 7.37 (0.11) | 50.53 (19.49) |
| **2** | 18.80 (3.99) | 39.83 (51.55) | 70.22 (13.05) | 2.06 (1.74) | 18.89 (11.17) | 35.48 (46.58) | 1.32 (1.37) | 7.36 (0.10) | 57.65 (19.69) |
| **3** | 17.70 (3.95) | 40.79 (56.98) | 71.71 (13.71) | 1.94 (1.99) | 17.87 (11.39) | 40.81 (51.26) | 1.24 (1.17) | 7.36 (0.11) | 58.72 (18.67) |
| **4** | 17.47 (4.81) | 41.90 (54.29) | 70.08 (14.16) | 2.45 (2.35) | 17.48 (12.13) | 38.36 (48.80) | 1.32 (1.31) | 7.39 (0.10) | 50.29 (19.45) |
| **5** | 17.42 (4.70) | 36.65 (45.83) | 69.93 (14.01) | 2.24 (2.03) | 18.66 (12.21) | 36.19 (47.40) | 1.29 (1.22) | 7.38 (0.11) | 47.86 (20.36) |

| | | | | | | | | | |
|---|---|---|---|---|---|---|---|---|---|
| **6** | 18.71 (4.12) | 39.26 (48.03) | 70.87 (13.14) | 2.09 (1.94) | 19.35 (11.05) | 35.65 (45.82) | 1.27 (1.27) | 7.36 (0.12) | 52.97 (19.96) |
| **7** | 18.90 (3.23) | 35.26 (51.15) | 68.91 (13.01) | 2.24 (1.93) | 17.62 (11.24) | 37.08 (46.18) | 1.39 (1.53) | 7.37 (0.12) | 65.65 (19.74) |
| **8** | 17.59 (4.29) | 39.93 (55.17) | 70.16 (14.02) | 2.15 (1.93) | 18.42 (11.72) | 37.93 (52.04) | 1.29 (1.28) | 7.38 (0.11) | 55.26 (17.80) |
| **9** | 17.41 (4.53) | 39.51 (50.49) | 70.48 (14.10) | 1.93 (1.57) | 17.71 (11.00) | 40.40 (53.36) | 1.26 (1.22) | 7.38 (0.10) | 52.47 (19.99) |

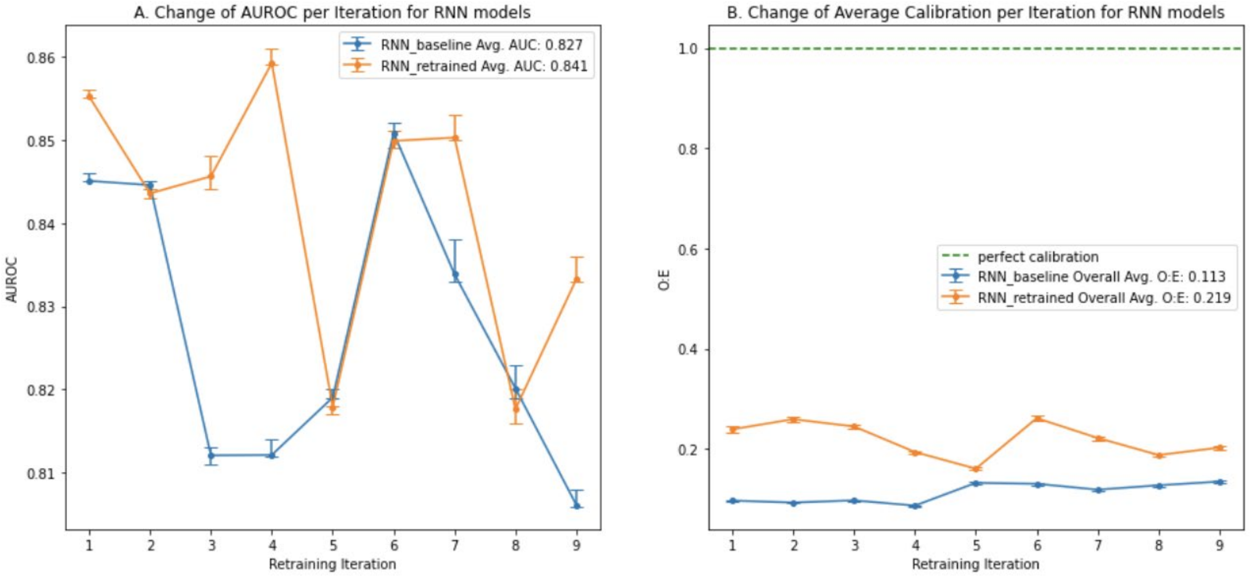


**Supplementary Figure 2.** Major event simulation. **A.** Changes of AUROC per iteration for RNN models. **B.** Changes to the average calibration per iteration for RNN models.

**Supplementary Table 2.** Covariate shift simulation. Distribution of the most important features. The values are in the format of mean (std). Iteration 0 is the baseline distribution.

| Iteration | RespRate (BPM) | HR (BPM) | WBC (1000/µL) | Temp (ºC) | RBC | Creatinine (mg/dL) | Lymphocytes (%) | Lactate (mmol/L) | SystolicBP (mmHg) | Monocytes (%) | BUN (mg/dL) | Age (years) |
|---|---|---|---|---|---|---|---|---|---|---|---|---|
| **0** | 17.81 (4.62) | 84.39 (17.58) | 9.92 (5.64) | 36.81 (0.44) | 3.84 (0.79) | 1.33 (1.69) | 18.30 (11.58) | 2.21 (1.95) | 124.68 (23.16) | 7.72 (3.46) | 22.41 (18.50) | 51.91 (20.13) |
| **1** | 18.00 (4.46) | 82.82 (17.18) | 10.13 (5.89) | 36.79 (0.44) | 3.97 (0.81) | 1.30 (1.29) | 18.63 (11.17) | 2.38 (2.23) | 125.02 (23.43) | 7.79 (3.30) | 21.98 (16.81) | 52.38 (19.87) |
| **2** | 18.63 (4.22) | 82.32 (17.29) | 10.21 (6.59) | 36.77 (0.46) | 3.94 (0.82) | 1.33 (1.28) | 19.25 (12.65) | 2.30 (2.34) | 126.12 (23.33) | 7.72 (3.14) | 23.73 (18.60) | 57.04 (19.78) |
| **3** | 17.88 (4.08) | 82.19 (16.06) | 10.22 (5.88) | 36.77 (0.39) | 3.91 (0.81) | 1.25 (1.23) | 17.34 (10.84) | 1.92 (1.92) | 127.75 (22.52) | 8.01 (3.27) | 22.79 (18.35) | 58.76 (18.84) |
| **4** | 17.47 (4.77) | 83.70 (17.37) | 9.95 (5.68) | 36.78 (0.36) | 3.77 (0.78) | 1.32 (1.25) | 17.83 (10.98) | 2.32 (2.29) | 124.04 (22.68) | 7.51 (3.33) | 22.85 (19.00) | 47.50 (19.67) |
| **5** | 17.79 (4.60) | 83.23 (17.60) | 10.38 (5.65) | 36.79 (0.42) | 3.91 (0.81) | 1.24 (1.12) | 19.11 (12.44) | 2.30 (2.38) | 124.75 (23.80) | 7.47 (3.40) | 22.38 (18.64) | 52.49 (19.76) |
| **6** | 19.26 (3.58) | 79.80 (17.12) | 9.67 (6.02) | 36.80 (0.48) | 4.10 (0.86) | 1.41 (1.54) | 18.03 (11.64) | 2.17 (1.77) | 128.04 (22.53) | 8.15 (3.63) | 24.37 (19.37) | 61.15 (20.77) |
| **7** | 18.09 (3.66) | 80.48 (16.88) | 10.10 (5.75) | 36.79 (0.45) | 3.80 (0.80) | 1.31 (1.35) | 17.65 (10.91) | 2.04 (1.70) | 126.33 (22.66) | 7.80 (3.65) | 22.98 (18.34) | 58.16 (19.52) |
| **8** | 17.56 (4.74) | 83.56 (17.27) | 10.45 (6.47) | 36.78 (0.38) | 3.80 (0.79) | 1.34 (1.36) | 18.08 (11.52) | 2.16 (2.02) | 124.49 (23.20) | 7.58 (3.27) | 23.08 (18.65) | 50.54 (19.03) |

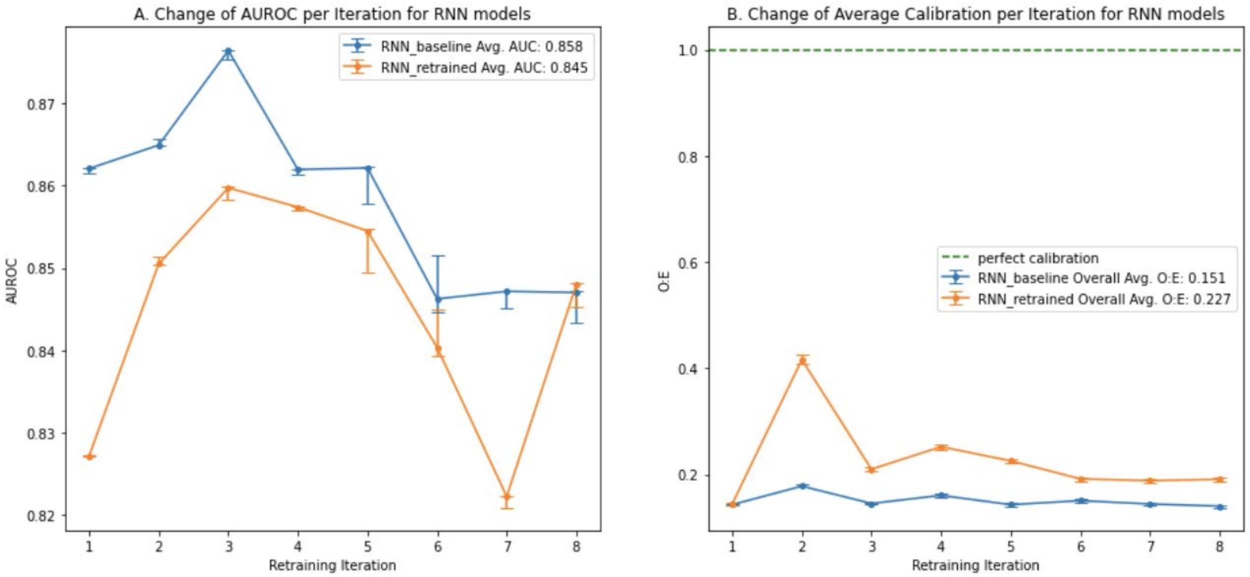


**Supplementary Figure 3.** Covariate shift simulation. **A.** Changes of AUROC per iteration for RNN models. **B.** Changes to the average calibration per iteration for RNN models.

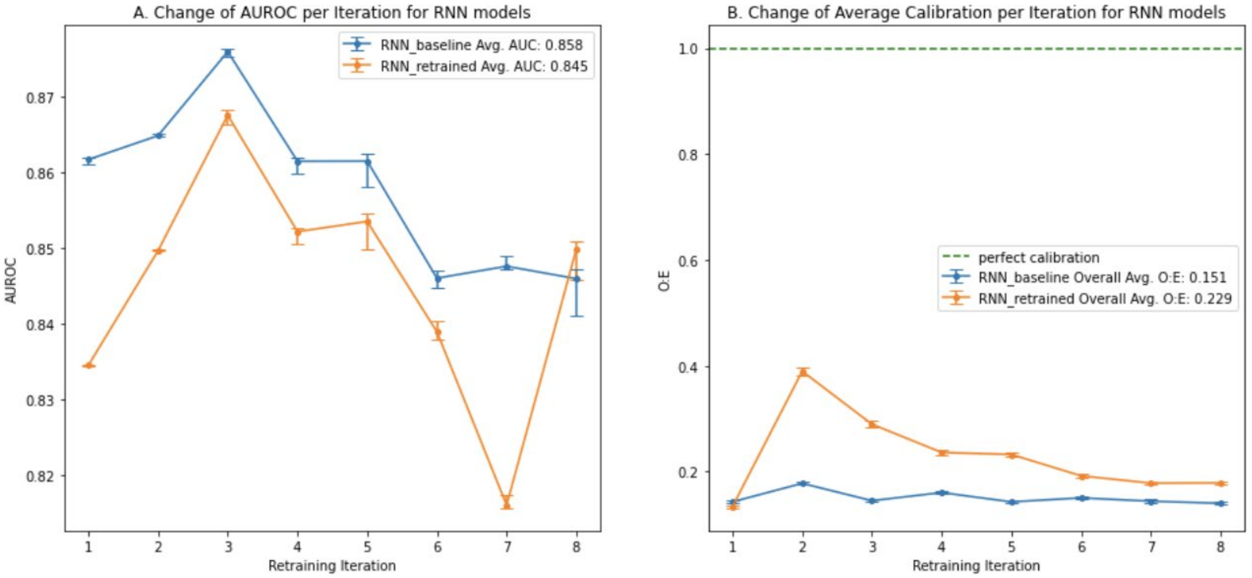


**Supplementary Figure 4.** Concept shift simulation, mixed labels. **A.** Changes of AUROC per iteration for RNN models. **B.** Changes to the average calibration per iteration for RNN models.
